# Genome-wide study on 72,298 Korean individuals in Korean biobank data for 76 traits identifies hundreds of novel loci

**DOI:** 10.1101/2022.02.23.22271389

**Authors:** Kisung Nam, Jangho Kim, Seunggeun Lee

## Abstract

Genome-wide association studies (GWAS) on diverse ancestry groups are lacking, resulting in deficits of genetic discoveries and polygenic scores. We conducted GWAS for 76 phenotypes in Korean biobank data, namely Korean Genome and Epidemiology Study (KoGES, n=72,298). Our analysis discovered 2,237 associated loci, including 117 novel associations, many of which replicated in Biobank Japan (BBJ) GWAS. We also applied several up-to-date methods for genetic association tests to increase the power, discovering additional associations that are not identified in simple case-control GWAS. We evaluated genetic pleiotropy to investigate genes associated with multiple traits. Following meta-analysis of 32 phenotypes between KoGES and Biobank Japan (BBJ), we further identified 379 novel associations and demonstrated the improved predictive performance of polygenic risk scores by using the meta-analysis results. The summary statistics of 76 KoGES GWAS phenotypes are publicly available, contributing to a better comprehension of the genetic architecture of the East Asian population.

## Introduction

Population-based biobanks such as UK Biobank^1,2^ and FinnGen^3^ facilitate genome-wide association studies (GWAS) in tens of thousands or even millions of samples across a large number of traits. These extensive resources helped identify numerous genetic associations and elucidate genetic components of complex traits^4,5^. Using the analysis results, genome-based prediction models have been built and successfully identified individuals with a high risk of disease. Despite the success, a major limitation of the current status of GWAS is the relative lack of non-European samples^6^. As rare variants in Europeans can have high minor allele frequencies (MAF) in other ancestry groups, the lack of non-European samples can limit further discoveries. In addition, it can cause health disparities if the use of genetic discovery in clinical practice is limited to European-ancestry individuals.^7^

We report a GWAS of 76 phenotypes in 72,298 Korean individuals from the Korean Genome and Epidemiology Study (KoGES), a large biobank conducted by the National Biobank of Korea. Recently, several studies have contributed to the catalog of genetic associations using East Asian biobanks such as Biobank Japan (BBJ)^8-10^ or Taiwan Biobank (TWB)^11^. By increasing the sample size and demographic diversity of East Asian GWAS samples, our analysis contributes to novel discoveries. Through the analysis of 14 binary diseases endpoints, 31 biomarkers, 23 dietary information, and 8 alcohol consumption phenotypes, we identified 2,237 associated loci for 43 phenotypes at the genome-wide significance level (*p* < 5 × 10^−8^). In order to fully use the information in the KoGES data, in addition to the mixed effect model for continuous, binary, and categorical phenotypes, we applied up-to-date analysis methods for survival GWAS^12^ and methods to incorporate family disease history^3^, and identified 19 additional significant associations. Among associations, 117 were novel, and more than 70 percent of novel associations with corresponding phenotypes and genetic variants in BBJ were replicated at a nominal p-value of 0.05. Many of the novel loci had very low MAF in Europeans, demonstrating power increment by utilizing samples from diverse ancestry groups.

To find East Asian-specific genetic associations, we conducted meta-analyses for 32 traits using KoGES and BBJ (n=251,000) GWAS results. We identified 379 novel loci for 25 traits, mostly in clinical biomarkers, and 85 percent of these loci were not identified in individual studies. We also constructed polygenic risk scores with meta-analyzed GWAS summary and showed that the PRS trained with the meta-analyzed KoGES and BBJ could have 20 percent larger R^2^ in the prediction of the trait-values in East Asian samples in UK Biobank compared to PRSs trained from BBJ GWAS only, demonstrating one potential utility of our analysis results. We publicly provide all GWAS summary statistics to broaden the understanding of the genetics basis of the East Asian population.

## Results

### Korean Genome and Epidemiology Study (KoGES)

KoGES, part of the National Biobank of Korea, is a prospective cohort study with a comprehensive range of phenotypic measures and biological samples such as DNA, serum, plasma, and urine collected on approximately 210,000 individuals. KoGES includes the community-based Ansan and Ansung study, the urban community-based health examinee study (HEXA), and the rural community-based cardiovascular disease association study (CAVAS). Each cohort has the baseline assessment and follow-up measurement thereafter, and we used the baseline measures only in this study (see Methods for detail). The table in Figure 1 describes the sample size and the mean age of KoGES data by cohort. Total 72,000 samples with Korean chip^13^ genotyped and imputed were used in our analysis.

**Figure 1.**
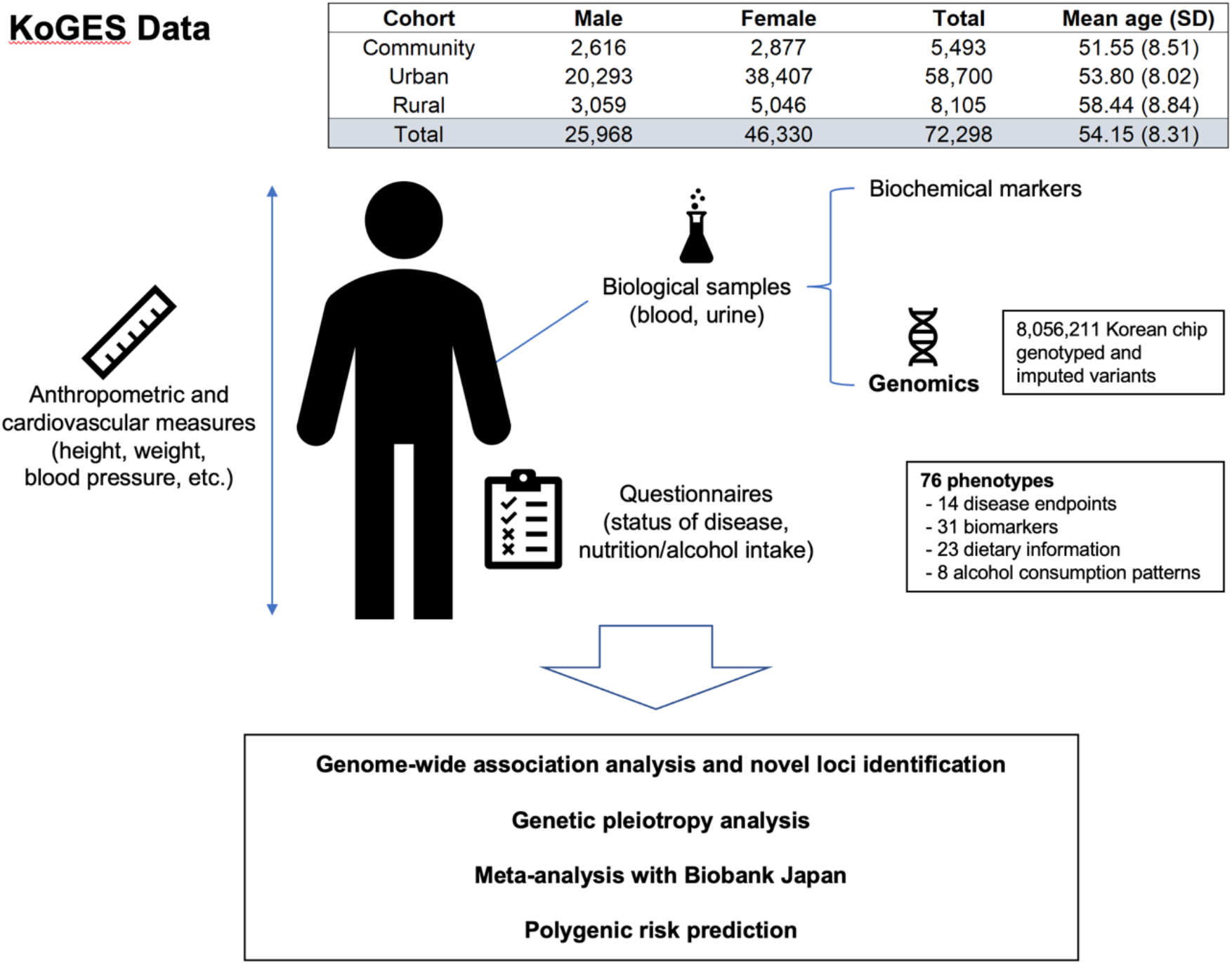
Overview of the KoGES data and our study KoGES data consists of three cohorts: community-based, the urban, and the rural cohort. Participants in KoGES are recruited from the national health examinee registry, age ≥ 40 at baseline. The sample size in the figure indicates the number of samples for which both genotype (after QC) and baseline assessment data exist.

### Genome-wide association analysis of 76 traits

The overview of our analysis is shown in Figure 1 and the studied traits are described in Supplementary Table 1. We analyzed 14 binary diseases endpoints, 31 biomarkers, 23 dietary information, and 8 traits about alcohol consumption patterns. Total 8,056,211 genotyped and imputed variants were used in our analysis. For continuous and binary traits, SAIGE^14^ was used to maximize power while controlling type I error. For ordinal categorical phenotypes, we applied a proportional odds logistic mixed effect model (POLMM)^15^. Total 2,218 loci for 43 traits satisfying a genome-wide significance threshold (*p* < 5 × 10^−8^) where significant clumped variants are identified using a window width of 5Mb and a linkage disequilibrium threshold of *R*^2^ = 0.1 (Supplementary Table 2). The estimated false discovery rate (FDR) is 0.0017 (FDR = 76 (# of traits) × 10^6^ (# of independent Loci) × 5 × 10^−8^ (genome-wide significance threshold) / 2218 (# of significant loci)). We performed linkage disequilibrium score regression (LDSC)^16,17^ to estimate heritability and genetic correlation (Supplementary Table 3). There were no significant confounding biases as the mean LDSC intercept values were 1.0139 for all 76 traits. As expected, height had the largest heritability (*h*^2^ = 0.400), followed by weight (*h*^2^ = 0.270) and blood platelet count (*h*^2^ = 0.265). The estimated heritabilities were similar to those of UKBB and BBJ (*h*^2^ = 0.402 and 0.386 for height, respectively). We additionally computed pairwise genetic correlations to discover the genetic relationship between phenotypes and represented them as a heatmap (Supplementary Figure 1). To avoid false positive findings, a genetic correlation was treated as zero when the p-value was greater than 0.05. We observed several phenotype clusters with high genetic correlations. 23 phenotypes related to nutrition intake form the largest cluster. In addition, we identified sets of closely related phenotypes such as (1) liver-related biochemical markers (aspartate transaminase, gamma-glutamyl transpeptidase, and alanine aminotransferase), (2) cardiovascular phenotypes (systolic blood pressure, diastolic blood pressure, and hypertension), and (3) hematological traits (hemoglobin, hematocrit, white blood cell count and red blood cell count).

Using the first onset age, we conducted survival analysis for 14 disease endpoints using SPACox, a method using Cox proportional hazards regression model. We replicated 15 significant associations for 3 traits that were not significant in case-control phenotype analysis. Incidence plots show that these loci influence the disease prevalence (Supplementary Figure 2). In addition, the association signals identified in case-control GWAS became more significant when applying survival analysis. (Supplementary Table 4).

By incorporating the family disease history, the association test power can be improved. We used the family disease history information of disease endpoints with TAPE^18^. In two types of cancer: gastric cancer and gallbladder cancer, we identified additional 4 independent association signals that were not detected in the analysis without family disease history. We calculated the mean value of the TAPE-adjusted phenotypes by the genotype of the top SNP of each locus (Supplementary Table 5). As the number of minor alleles increases, we observe that the TAPE-adjusted phenotypes monotonically increase or decrease.

### Genetic pleiotropy analysis

Since our analysis results show numerous associations, we investigated pleiotropy. To evaluate it at a gene level, we first mapped the most significant variant in each associated locus to a gene using FUMA^19^ and then counted the number of phenotypes associated (Supplementary Table 6). Overall, 826 genes had more than 2 associations. Many genes in chromosome 12 showed high levels of pleiotropy. Two neighboring genes in chromosome 12, ERP29 (12:112,451,230-112,461,253; GRCh37) and NAA25 (12: 112,464,493-112,546,587; GRCh37), were the most pleiotropic genes with 28 associated phenotypes, and then 8 genes, including ALDH2, with 27 associated phenotypes (Figure 2). Except for genes in chromosome 12, GCKR in chromosome 2, associated with 11 phenotypes, was the most pleiotropic. GCKR encodes glucokinase regulatory protein and is related to many phenotypes affected by glucose metabolism, such as fasting glucose and insulin measurement^20^. This result is consistent with the studies on other biobank data such as BBJ and UKB^10^.

**Figure 2.**
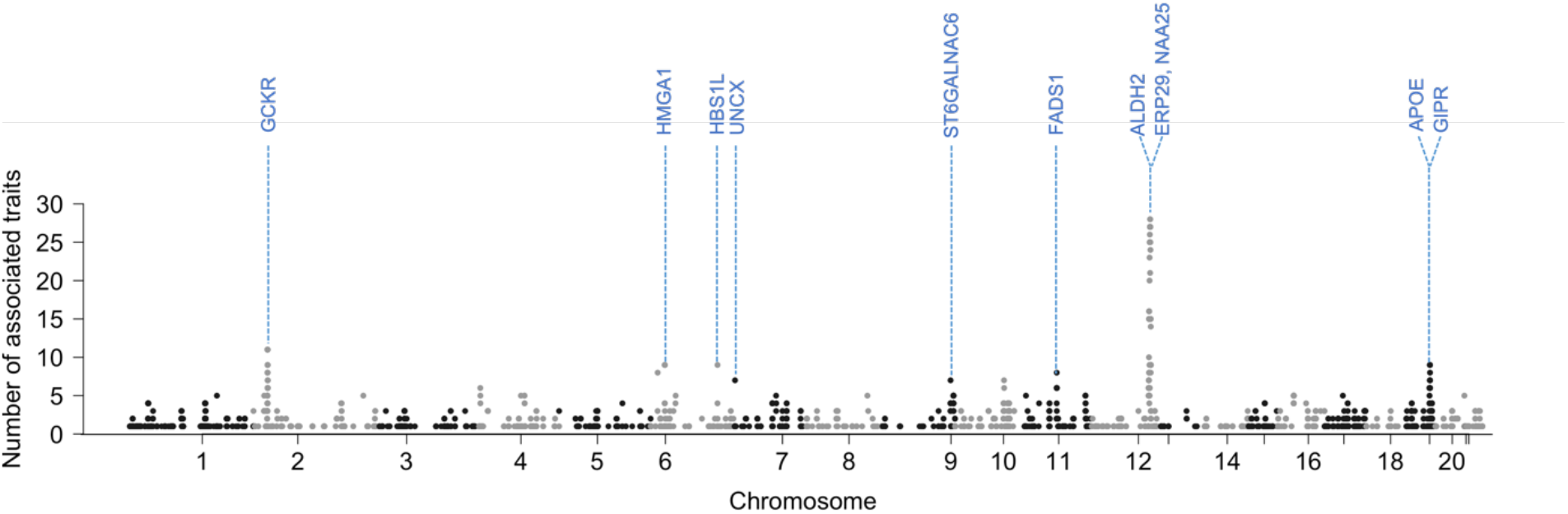
Manhattan plot with the y-axis being the number of significantly associated (p < 5 × 10^−8^) traits per gene for 76 traits in KoGES data. To avoid double-counting the associations in the same phenotypes, results from SPACox and TAPE were excluded when counting the number of significant associations.

The number of associated traits per variant is also used to quantify the degree of pleiotropy. 131 variants in chromosome 12 were associated with more than 10 traits. SNP rs11066132 and rs116873087, intron variants in NAA25, were the most pleiotropic variants (23 traits). Except for variants in chromosome 12, rs1260326, a missense variant at the GCKR locus (and 2:27,745,764), was the most pleiotropic variant (11 traits).

### Novel associations and replications

We identified 117 novel associations for 28 traits among the significant associations (Supplementary Table 7). We defined the association as novel if the association is not reported in the GWAS catalog and the p-value is not genome-wide significant (p-value < 5 × 10^−8^) in the BBJ GWAS (see Method). Among 117 novel associations, 53 top SNPs for 18 phenotypes were present in BBJ, and 38 SNPs (71.7%) out of 53 had BBJ p-value < 0.05. Many of the corresponding variants have low minor allele frequencies in the European population (Supplementary Figure 3). For example, rs939955, an intergenic variant between CYP3A4-CYP3A7, was associated with triglycerides level (*p* = 2.47 × 10^−9^, BBJ p-value = 1.2 × 10^−4^). The MAF of rs939955 in KoGES was 0.22, but it was very rare among Europeans (MAF_EUR_=0.002). Both CYP3A4 and CYP3A7 belong to Cytochrome P450 (CYP) superfamily and are well known for drug metabolism. A variant rs1314013, at the ZEB1 locus, was associated with weight (*p* = 7.19 × 10^−11^, BBJ p-value = 2.7 × 10^−4^, MAF_KoGES_=0.17, MAF_EUR_=0.04). In an experiment on mice, the zinc finger E-box binding homeobox (ZEB1) transcription factor was a repressor of adiposity^21^. A variant rs118190473, at ANXA3, was associated with HDL cholesterol level (*p* = 4.49 × 10^−8^, p-value in BBJ = 7.0 × 10^−6^, MAF_KoGES_=0.155, MAF_EUR_=0.005). Since adipocyte differentiation and lipid accumulation is the potential function of Annexin A3 (ANXA3)^22,23^, our result may provide a link between HDL level and the ANXA3 locus. We identified rs9921399, an intron variant at CES1, was associated with LDL cholesterol (*p* = 1.37 × 10^−9^, BBJ p-value = 5.5 × 10^−5^, MAF_KoGES_=0.45, MAF_EUR_=0.23). Although this locus has not been demonstrated in GWAS, it is known that LDL cholesterol levels were reduced in carboxylesterase 1 (CES1) deficient mice.^24^ We compared effect sizes and 95% confidence interval for those 4 loci and drew locus zoom plots (Supplementary Figure 4).

### Meta-analysis with Biobank Japan (BBJ)

To identify genetic associations in the East Asian population, we conducted a meta-analysis for 9 disease endpoints and 23 biomarkers for KoGES together with Biobank Japan (BBJ) and identified 289 genome-wide significant associations for diseases endpoints and 6,197 significant associations for biomarkers (Supplementary Table 8). Figure 3 represents the number of associations identified in the meta-analysis across KoGES and BBJ. As expected, meta-analysis substantially increased the number of significant associations. Of the total 6,486 associated locus-trait pairs, 1,677 (25.8%) were newly identified by the meta-analysis. For example, alanine aminotransferase GWAS identified 26 loci in KoGES and 52 in BBJ, yet 124 loci were significant in the meta-analysis. Among the identified associations, 379 (2 disease-associated and 377 biomarker-associated) were novel and 321 novel associations were not significant in individual GWAS of KoGES or BBJ.

**Figure 3.**
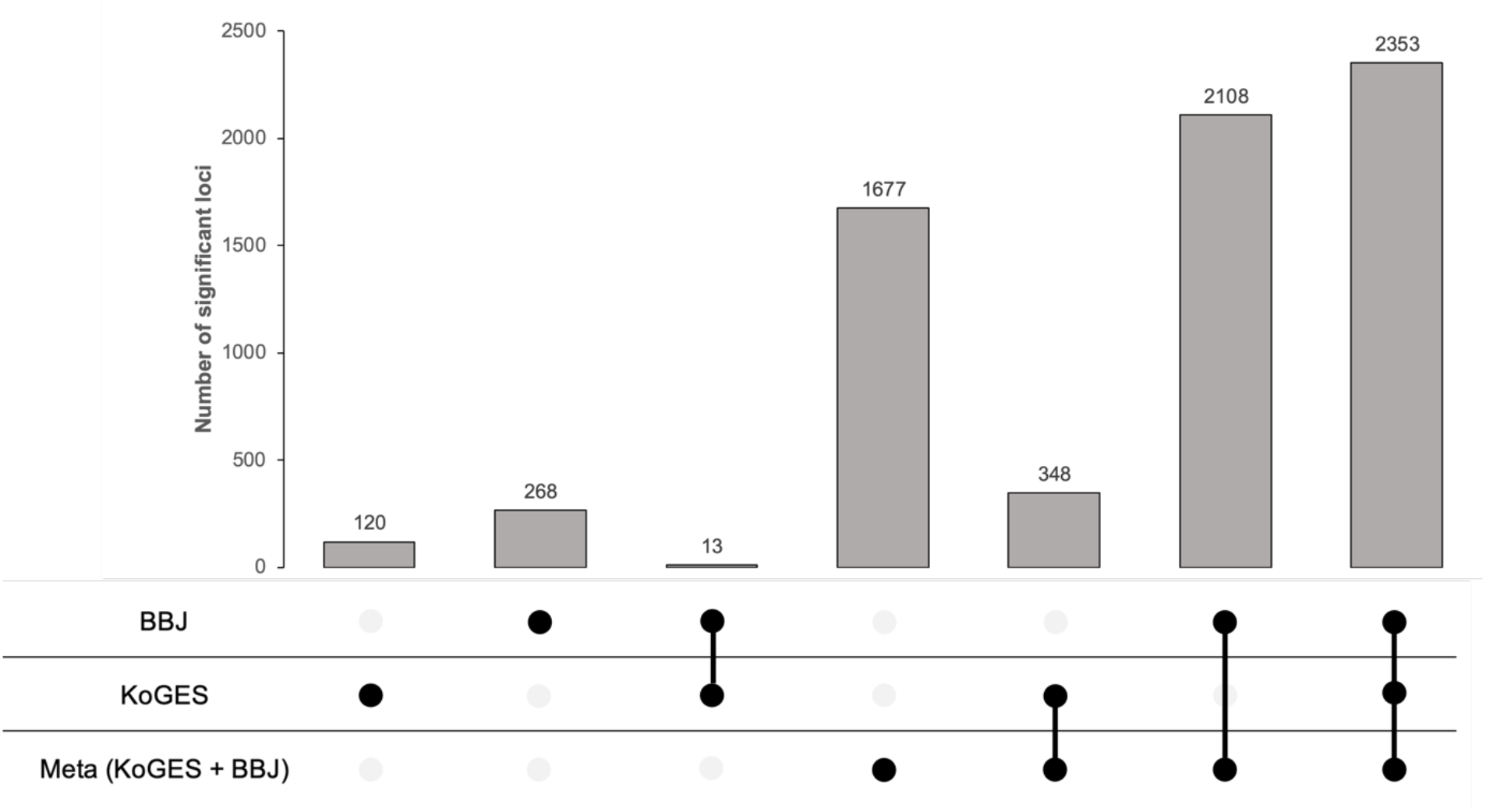
Number of significant associations identified in the KoGES and BBJ meta-analysis. Black dots indicate significance in the analysis.

### Polygenic Risk Score Improvement

We calculated polygenic risk scores (PRS) based on the meta-analysis GWAS results across KoGES and BBJ and compared them with the BBJ-based PRS model. Using PRS-CS^25^, we trained the PRS model for systolic blood pressure (SBP), diastolic blood pressure (DBP), high-density lipoprotein cholesterol (HDLC), low-density lipoprotein cholesterol (LDLC), and triglycerides (TG). To estimate unbiased prediction performance, we used East Asian samples in UK Biobank as the test samples. For all 5 phenotypes we tested, PRS based on the meta-analysis provides better predictive performance, in terms of R-squared, compared to BBJ-based PRS, in all models that adjusted for age and sex as covariates (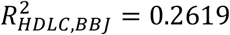 and 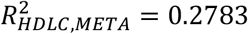), models adjusted for sex only (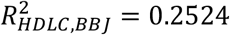 and 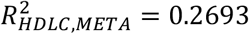) and that without adjustment (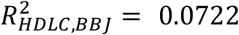 and 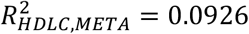). The entire analysis result is described in Supplementary Table 9. This result demonstrates that the meta-analysis including our KoGES GWAS can contribute to the improvement of risk prediction for East Asians.

## Discussion

In this paper, we carried out GWAS for 76 phenotypes in 72,000 Korean samples and identified a large number of significant associations. Our analysis found 117 novel associations previously unknown, and many of them were replicated in BBJ. We also performed pleiotropy analysis and illustrated that the most pleiotropic regions such as a region of chromosome 12 including ERP29, NAA25, and ALDH2, and a chromosome 2 region including GCKR. Through the meta-analysis with BBJ, we further identified a large number of significant associations. Since most of the novel associations in the meta-analysis with BBJ were not identified in individual GWAS, our study contributes to increasing the effective sample size. We additionally compared the prediction models based on the meta-analysis results and BBJ summary statistics. We demonstrated that a model based on the meta-analysis across KoGES and BBJ has better prediction performance when predicting trait values for East Asian samples in UK Biobank.

Biobanks collect additional disease information from either survey or electronic records, such as time to disease onset and family disease history. In addition to genetic association analysis of continuous, binary, and categorical phenotypes, we applied survival analysis (SPACox) and model with family history (TAPE) to utilize time-to-onset age and family history. These methods found 19 more significant loci in 5 traits; all of them were previously known, validating the approach. In addition, many of the significant loci p-values were improved. Our analysis demonstrates that time-to-onset and family history are valuable data and can help to identify true associations.

In our pleiotropy analysis, many genes in chromosome 12 were highly pleiotropic and many of the associated phenotypes were alcohol consumption-related. Based on this, it is reasonable to assume that ALDH2 association with alcohol drive the signals. The variant rs671 in ALDH2 had a significant negative effect on alcohol consumption, hence, affected many alcohol consumption phenotypes and alcohol-related phenotypes such as blood pressure and cholesterol level. Interestingly, ERP29 and NAA25 had an additional association with thyroid disease. NAA25 is known for its association with hypothyroidism^26^, and both ERP29 and NAA25 genes are linked to several traits, including blood pressure and alcohol-related traits^27^

There are several limitations in our study. First, the disease status phenotypes in KoGES are collected through a self-reported survey and have not been verified by an expert diagnosis. Second, the nutrition intake data in KoGES is derived from a food frequency questionnaire (FFQ) involving 103 foods (See methods), which can have errors. Third, since there is no information on medication in KoGES data, calibration of several continuous phenotypes, such as blood pressure, was not feasible. Despite the limitations, our study is the only existing research that analyzed a large number of phenotypes in the Korean population.

To better predict and prevent complex diseases, we need large GWAS in diverse populations. Ideally, GWAS results should be publicly available for meta-analysis and downstream analysis, including novel association identification and replication, PRS calculation, and Mendelian Randomization. All our GWAS and meta-analysis results are publicly available on a PheWeb^28^ website with interactive visualization of Manhattan, Q-Q, locus zoom plots. By providing East Asian GWAS on many phenotypes, our results will contribute to elucidating the genetic architecture of complex traits.

## Methods

### KoGES data

We used measures of the baseline recruitment, and only genotyped samples that met the following exclusion criteria were used: low call rate (< 97%), excessive heterozygosity, excessive singletons, gender discrepancy, and cryptic first-degree relatives. SNPs with low HWE p-value (< 10^−6^) or low call rate (< 95%) were excluded. After quality control, data were phased using Eagle v2.3 and imputed using IMPUTE4 with 1000 Genomes Project Phase 3 data and Korean reference genome as a reference panel. Variants with imputation quality score (IQS) < 0.8 and MAF < 1% were excluded after imputation. We analyzed 8,056,211 variants in total after these processes.

Anthropometric and clinical measurements in KoGES were obtained by physical examinations and clinical investigations, and the disease status of the participants and family members were collected by the interview. Nutritional intake data were calculated based on a categorical food frequency questionnaire (FFQ) involving 103 foods. Detailed methods for the measurements, interviews, and questionnaires are described elsewhere.^29,30^

### GWAS of KoGES data

For both binary and quantitative traits, we conducted GWAS using a linear mixed model implemented in SAIGE (version 0.44.5) in order to maximize power while controlling type I error for case-control imbalance. In step 1, we used 327,540 variants with Imputation Quality Score (IQS) = 1 to obtain the genetic relationship matrix (GRM). We used the top 10 principal components (PC), age, sex, and adjustment of assessment details such as cohort, year of examination, as covariates in step 1. For quantitative traits, we applied rank-based inverse normal transformation and leave-one-chromosome-out (LOCO) scheme to remove the proximal contamination. For ordinal categorical traits, we used a proportional odds logistic mixed model (POLMM) to model the nature of ordinal categorical phenotypes. POLMM is also known to be robust for imbalanced phenotypic distributions, while linear mixed models do not control type I error rate well. After GWAS, we conducted clumping analysis using PLINK2^31^ for the variants with p-values less than 5 × 10^−8^, window size of 5Mb, and linkage disequilibrium threshold R^2^ of 0.1 to count independent genome-wide significant loci. We carried out LD score regression using ldsc (version 1.0.1) to estimate SNP-based heritability, confounding bias, and genetic correlation. We reported the results for all 76 studied traits (Supplementary Table 2).

We performed survival analysis for 14 disease endpoints with SPACox (version 0.1.2), a Cox proportional hazards regression model with saddlepoint approximation, using the first onset age in KoGES. We used the same covariates (top 10 PCs, age, sex, and batch effect).

In KoGES, family disease history data collected by the survey are available. This data indicates whether a family member (separated in paternal, maternal, and siblings) has ever been diagnosed with the disease. With these family disease histories in our data, we conducted association analysis using TAPE (version 0.2.1). We first adjusted phenotypes (disease status) using three types of family history of disease: paternal, maternal, and siblings for 12 disease endpoints. In this study, we adjusted phenotypes assuming *ρ*, a pre-specified constant indicating the increase in latent disease risk, is equal to 0.5.

### Gene-level Genetic pleiotropy analysis

We measured the degree of pleiotropy by counting the number of associated traits (out of 76 traits) per gene and per variant for KoGES GWASs. We excluded the results from survival analysis (SPACox) and model with family history (TAPE) when counting the number of associated traits. For gene-level pleiotropy, we used the SNP2GENE function of FUMA^19^ to map SNPs in GWAS results to a gene with 1000 Genome Phase 3 EAS as a reference panel. All parameters used in gene mapping are default values provided by FUMA. For variant-level pleiotropy, we counted the number of associated traits satisfying a genome-wide significance threshold (*p* < 5 × 10^−8^).

### Novel association identification

We searched for existing associations using the GWAS catalog^32^ within ±500kb from the lead variant to regard an associated locus as novel. In order to screen for variants that have been previously reported, we first used gwasrapidd^33^ R package to access the representational state transfer (REST) application programming interface (API) of the GWAS catalog. To map traits reported under different names of traits, we used experimental factor ontology (EFO) traits, and we again exhaustively searched for existing reports of association in the same or similar phenotypes. For example, variants that have been reported for blood-pressure-related traits were excluded from the novel loci for hypertension. Since the results by BBJ^10^ were not listed in the GWAS catalog at the time of evaluation for the novel association, we additionally excluded variants that were genome-wide significant in BBJ.

### Meta-analysis with Biobank Japan

Summary statistics of BBJ were downloaded from the Biobank Japan PheWeb website (https://pheweb.jp/). 9 disease endpoints and 23 biomarker phenotypes were presented in both KoGES and BBJ, and a total of 6,907,490 variants were overlapped in these two studies. The z-score based meta-analysis method was used to calculate p-values and we used the inverse variance method to obtain the effect sizes for risk prediction. To identify the novel loci, we applied the same criteria as mentioned in the previous section.

### PRS evaluation

We calculated polygenic risk scores (PRS) using PRS-CS (version released on June 4, 2021). For both BBJ-based and meta-analysis-based models, we used East Asian (EAS) in 1000 Genomes Project phase 3 samples as the LD reference panel. All parameters used in our analysis are default values provided by PRS-CS software, and we did not specify the global shrinkage parameter phi to use a fully Bayesian approach. When training the PRS models, we used summary statistics after filtering variants that exist in all UK Biobank, KoGES, and BBJ. We additionally restricted variants in HapMap3 as in Prive et al^34^, so a total of 900,746 variants were used. The effect sizes of the meta-analysis were calculated from inverse variance methods described in the meta-analysis part.

## Data Availability

All data produced are available online at

https://koges.leelabsg.org

## Data availability

The summary statistics, Manhattan plots, and quantile-quantile plots for 76 phenotypes in the KoGES, 32 phenotypes in the meta-analysis are available for public download at https://koges.leelabsg.org. BBJ summary statistics used in this study were downloaded from the following: https://pheweb.jp/.

## Code availability

We used publicly available software for the analyses.

SAIGE: https://github.com/weizhouUMICH/SAIGE

POLMM: https://github.com/WenjianBI/POLMM

SPACox: https://github.com/WenjianBI/SPACox

TAPE: https://github.com/styvon/TAPE

LDSC: https://github.com/bulik/ldsc

FUMA: https://fuma.ctglab.nl/

PRS-CS: https://github.com/getian107/PRScs

PLINK2: https://www.cog-genomics.org/plink/2.0/

gwasrapidd: https://github.com/ramiromagno/gwasrapidd

## Acknowledgements

This research was supported by Brain Pool Plus (BP+, Brain Pool+) Program through the National Research Foundation of Korea (NRF) funded by the Ministry of Science and ICT (2020H1D3A2A03100666). Data in this study were from the Korean Genome and Epidemiology Study (KoGES; 4851-302), National Research Institute of Health, Centers for Disease Control and Prevention, Ministry for Health and Welfare, Republic of Korea. UK Biobank data were accessed under the accession number UKB: 45227. We thank Dr. Cristen Willer for the constructive comments and suggestions.

## Author contributions

K.N. and S.L. designed the experiments. K.N., J.K., and S.L. analyzed the KoGES data. K.N. and S.L. wrote the manuscript. All authors reviewed and approved the final version of the manuscript.

