## Supplementary Figures for "Genome-wide study on 72,298 Korean individuals in Korean biobank data for 76 traits identifies hundreds of novel loci"

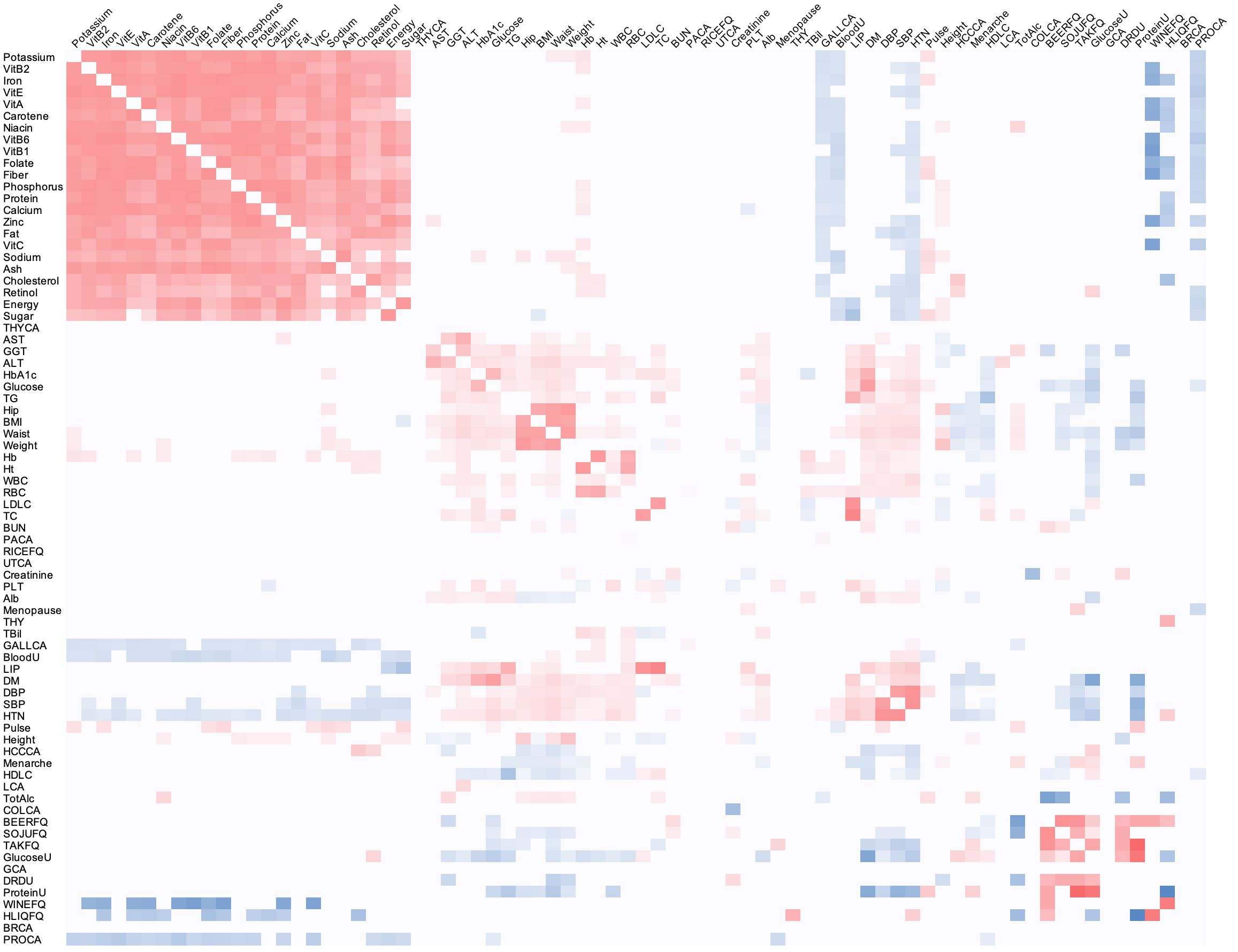

**Supplementary Figure 1.** Heatmap for pairwise genetic correlations. To reduce false positives, genetic correlation was treated as zero when the corresponding p-value is greater than 0.05.

(a)

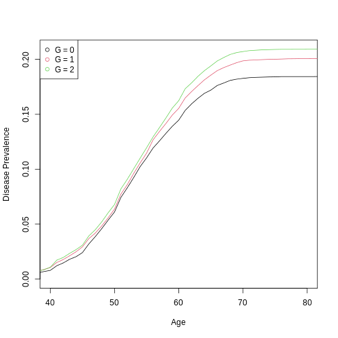

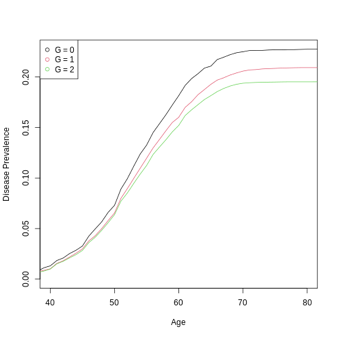

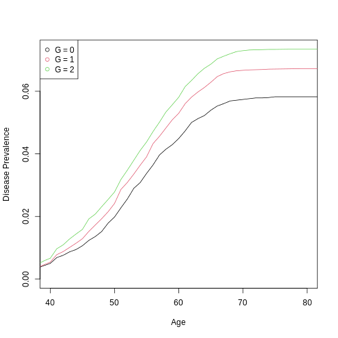

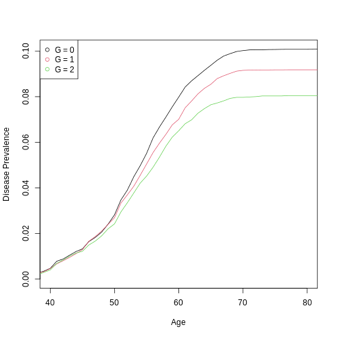

(b)

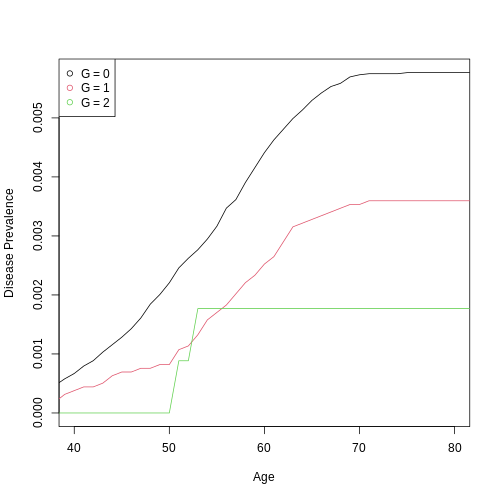

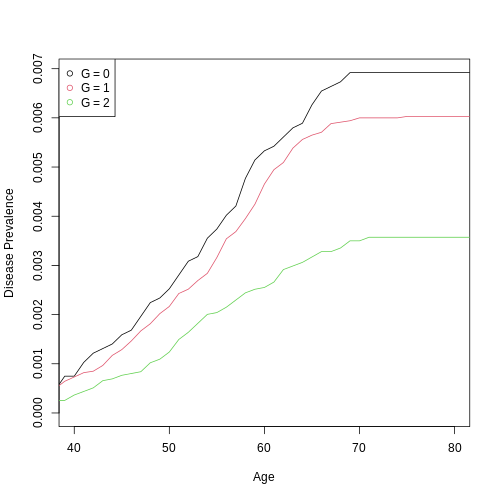

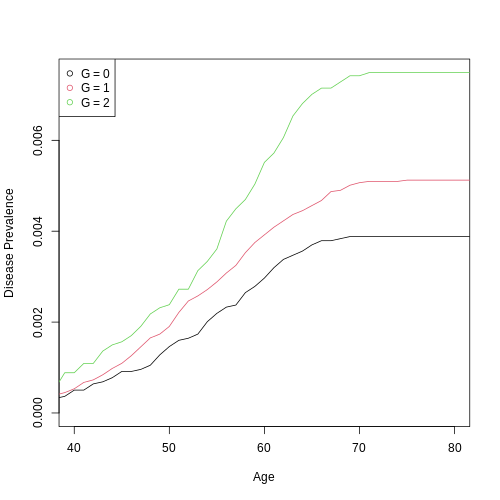

**Supplementary Figure 2.** (a) Incidence plot of three disease prevalence by genotype of significant variant. First two plots show the disease prevalence of hypertension (rs71037444 and rs113628671, respectively), and next two plots are of diabetes (rs201174461) and hyperlipidemia (chr16:72,003,267), respectively. (b) Incidence plot of gastric cancer by genotype of most significant loci. Each plot demonstrates the disease prevalence according to the genotype of rs760077, rs2978977, rs866605438, respectively.

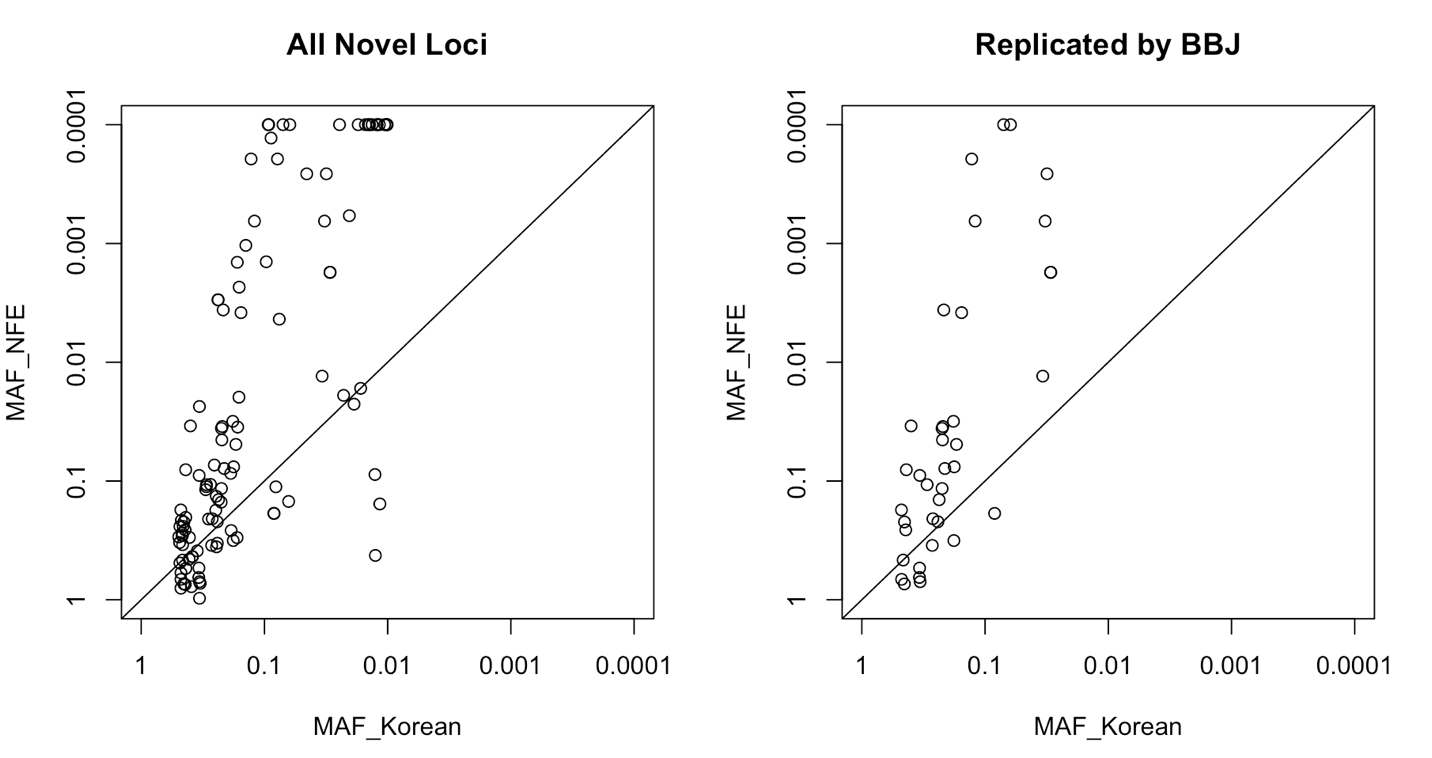

**Supplementary Figure 3.** Comparison of minor allele frequencies (MAF) in Korean and non-Finnish European (NFE) for the top variants of 117 novel associations. MAF estimates in gnomAD were used for NFE. When the MAF is less than 0.0001 or unavailable for NFE, we regarded them as 0.0001 in the plot. 84 variants (71.8%) had lower MAF in NFE than in Korean among all novel loci. After filtering variants replicated by BBJ (p-value less than 0.05 in BBJ), 29 variants (76.3%) had lower MAF in NFE.

(a)

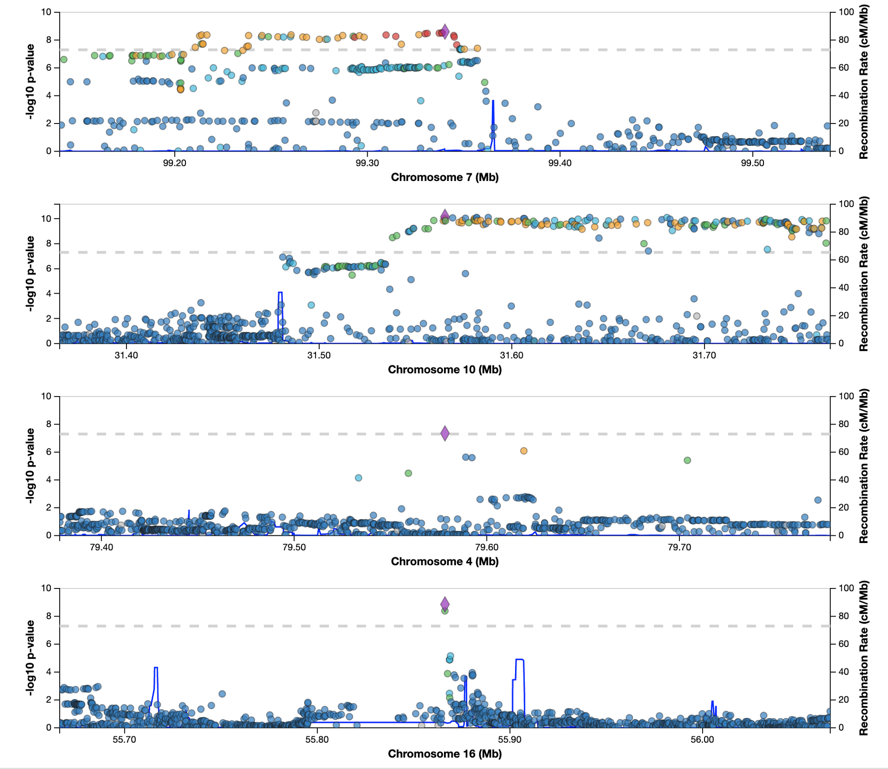

(b)

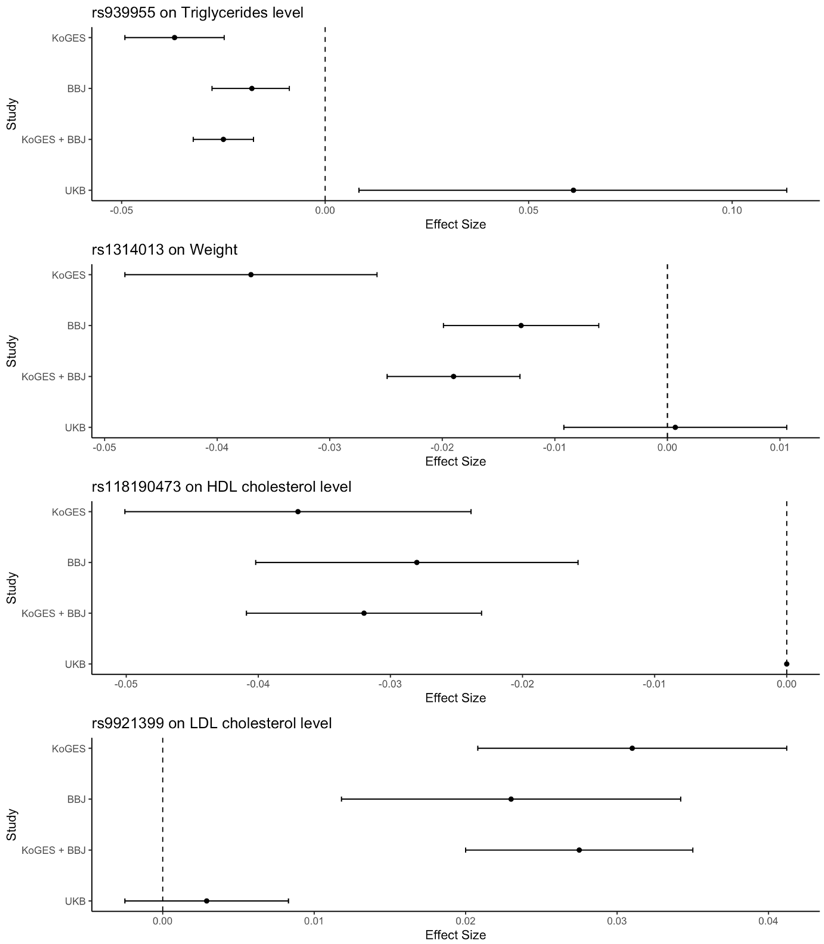

**Supplementary Figure 4.** (a) Locus zoom plots for selected novel loci. (b) Forest plots for effect sizes of top variant of selected novel loci. Each plot represents (1) rs939955 on triglycerides level, (2) rs1314013 on weight, (3) rs118190473 on HDL cholesterol level and (4) rs9921399 on LDL cholesterol level, respectively.
